# Harnessing the Eccrine Sweat Glands for the Management of Interdialytic Weight Gain – A Pilot Study

**DOI:** 10.1101/2024.04.15.24304270

**Authors:** Zaher A. Armaly, Yaacov Nitzan, Gil Chernin, Doron Aronson

## Abstract

**Background:** Hemodialysis patients are susceptible to excess volume accumulation, particularly over the 2-day interval (long interdialytic gap), resulting in higher interdialytic weight gain (IDWG).

**Methods:** We employed a novel device designed to enhance fluid and salt loss by activating of the eccrine sweat glands to treat IDWG. Patients eligible for the study were undergoing regular hemodialysis for ≥3 months, without residual renal function, and with IDWG (as a percentage of IDWG/dry body weight) ≥2.5%. Treatments were administered at the patient’s home. The primary performance endpoint was differences in weight gain over long interdialytic period with the device compared to the control period. Secondary exploratory endpoints included the need for ultrafiltration (UF) rate > 10 mL/kg/h in the post-interval dialysis.

**Results:** Five patients were enrolled into the pilot study (age range 26 to 69 years, 3 women). The hourly mean weight loss from sweat was 186 ± 45 gr/h. The average procedure length was 4.5 hours with an average fluid loss of 899 ± 283 gr per procedure. The average least-squares mean absolute difference between the control and treatment periods was -2.0%; (95% confidence interval [CI], -2.9% to -1.2%, P<0001). The reduction in IDWG was associated a reduction in UF rates, with a least-squares mean difference of -3.2 mL/Kg/min [95% CI -4.6 to -1.8] between the control and treatment periods (P<0.001), resulting in a safe UF rate (<10 mL/Kg/min) in all post-treatment sessions.

**Conclusion:** Enhancing sweat rate mitigated IDWG in hemodialysis patients. Together with trice weekly HD, this therapy more closely approximates the capacity of the native kidney to regulate extracellular volume and solute composition, similar to patients with substantial residual kidney function.

## Introduction

Volume overload is prevalent among patients undergoing hemodialysis and is associated with diminished quality of life and adverse outcomes including systemic hypertension, left ventricular hypertrophy, hospitalizations and mortality.^1, 2^ Hemodialysis patients are susceptible to excess volume accumulation, particularly over the 2-day interval (long interdialytic gap), resulting in higher interdialytic weight gain (IDWG).^1-5^

IDWG is closely tied to elevated ultrafiltration rates. Rapid ultrafiltration rate following the long interdialytic gap, coupled with excessive IDWG, promotes non-physiological fluid shifts and hemodynamic instability, pre-syncope/syncopal events, and symptoms such as cramping and postdialysis fatigue.^6^ The downstream consequences of ischemic organ injury associated with high ultrafiltration rates include rapid loss of residual kidney function,^7^ cerebral ischemia, brain white matter damage and dementia,^8, 9^ myocardial stunning with maladaptive cardiac structural changes,^10^ arrhythmia, cardiac sudden death, mesenteric ischemia and gut related systemic endotoxemia^10, 11^ and overall mortality.^12^ Thus, there exists a narrow therapeutic window to avoids both excessive IDWG and high UF rates, along with the resulting complications of both volume depletion and overload.^4^

Interest in the use of sweat glands to treat renal diseases has been present in the nephrology literature for decades. The eccrine sweat glands can produce large amounts of fluid in addition to other constituents including sodium, chloride, potassium, phosphorous and ammonium.^13^ We have recently shown that enhancing sweat rate was safe and resulted in a clinically meaningful fluid removal rates in patients with heart failure.^14^ In this pilot study, we explore this concept for the treatment of IDWG in hemodialysis patients.

## Methods

We employed the AquaPass system (AquaPass Medical Ltd., Shefayim, Israel), designed to enhance fluid and salt loss by activating of the eccrine sweat glands.^14^ The AquaPass system (AquaPass Medical Ltd., Shefayim, Israel) is designed to enhance fluid and salt loss via the eccrine sweat glands, has been previously described.^14, 15^ Briefly, the system is comprised of two main functional units: (1) a capsule and (2) heating units with controller. The capsule creates a homogeneous warm temperature environment around the lower part of the body leading to increased skin temperature that activates the eccrine glands and initiates perspiration. The sweat evaporates instantaneously, thus avoiding the awareness of perspiration by the patient and enabling long durations of treatments, if required.

The heating sub-unit controls temperature inside the capsule. The skin temperature is uniformly increased from 32-33□ to the range of 36–40□, where the slope of the relationship between temperature and sweat production is linear2-4 and discomfort or thermal injury does not occur.^16^ In a first-in-human study of stable heart failure patients, the median hourly weight loss induced by the device was 215 g/h (interquartile range, 165–285; range, 100–344 g/h).^14^

A user interface enables the operator to set the air flow rate and skin temperature within allowed range of values. During the treatment, measurements made by the sensors in the wearable and the calculated sweat rate are displayed.

With increased sweat rate, the majority of the fluid removed is from the interstitial compartment as interstitial fluid is the precursor fluid for sweat (in contrast to mobilization of fluid from the intravascular space with hemodialysis).

The study was conducted with the approval of the institutional review boards at the Nazareth Hospital EMMS, Nazareth, Israel. Written informed consent was obtained from all study participants. The protocol was registered at ClinicalTrials.gov (Unique identifier: NCT06358365).

Patients eligible for the study were undergoing regular hemodialysis for ≥3 months, without residual renal function (RKF), and with IDWG (as a percentage of IDWG/dry body weight) ≥2.5%. Primary safety endpoint was device related serious adverse events. The primary performance endpoint was differences in weight gain over long interdialytic period with the device compared to the control period. Secondary exploratory endpoints included the need for ultrafiltration rate > 10 mL/kg/h in the post-interval dialysis, hypotension in the post-interval dialysis session, systolic blood pressure pre-dialysis after the dialysis interval; Quality of life as assessed by the Kansas City Cardiac Questionnaire-12 (KCCQ-12); changes in hemoglobin, BUN, and electrolytes, and changes in biomarkers of congestion (NT-proBNP).

The study was conducted with the approval of the institutional review boards at the Nazareth Hospital EMMS, Nazareth, Israel. Written informed consent was obtained from all study participants before the study procedures. The study flow is depicted in Figure 1. Patients entered a 3-week observation period, followed by a 4-week treatment period. In addition to the regular planned hemodialysis sessions, each patient underwent 2 weekly procedures (∼4h) with the AquaPass device during the weekend at the patient’s home. We used repeated measurements ANOVA to determine differences in IDWG and UF rates over time, with weighted contrast between the first 3 and last 4 repeated measurements. All other statistical comparisons were exploratory.

**Figure 1:**
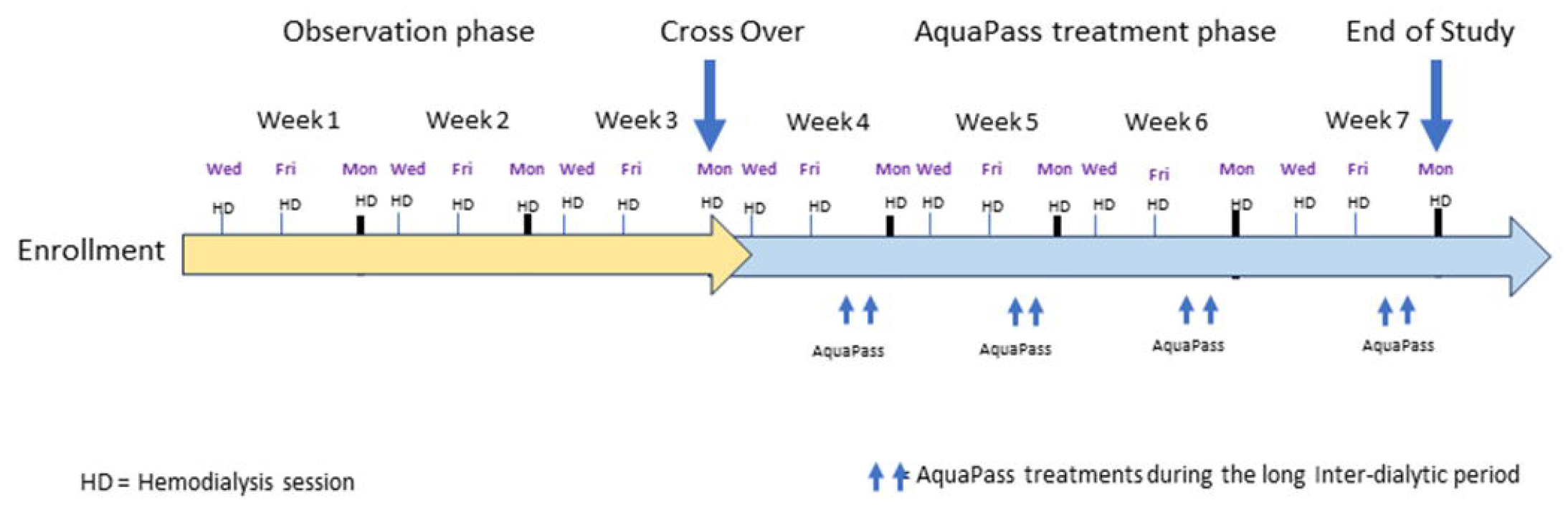
Study Schedule

## Results

Five patients were enrolled into the pilot study (age range 26 to 69 years, 3 women). The mean IDWG during the control period was 3.4 ± 1.3%. During the home therapy, the hourly mean weight loss from sweat was 186 ± 45 gr/h. The average procedure length was 4.5 hours with an average fluid loss of 899 ± 283 gr per procedure. There were no procedure-related adverse events.

During the treatment period patients had lower IDWG than in the control period (Figure 2A). The average least-squares mean absolute difference between the periods was -2.0%; (95% confidence interval [CI], -2.9% to -1.2%, *P*<0001). The reduction in IDWG was associated a reduction in UF rates, with a least-squares mean difference of -3.2 mL/Kg/min [95% CI -4.6 to -1.8] between the control and treatment periods (*P*<0.001), resulting in a safe UF rate in all post-treatment sessions (Figure 2B). AquaPass therapy was also associated with a reduction in potassium levels (least-squares mean difference -0.28 mmol/L [95 CI -0.56 to - 0.04, *P*=0.025]. We also observed an increase in hemoglobin and calcium levels and a reduction in NT-proBNP (Table 1). The KCCQ-12 score increased from a baseline of 67 ± 12 to 86 ± 11 at the end of the study (Figure 3). After the 4^th^ weekend treatments, the scheduled hemodialysis was deferred by an additional 24h in all patients.

**Table 1:** Laboratory tests in the control and treatment periods.

| Test | Least Square Mean Difference of Treatment vs. | *P* |
| --- | --- | --- |
|  | Control (95% CI) | value* |
| Haemoglobin (g/dl) | 0.5 [0.3 to 0.8] | <0.001 |
| BUN (mg/dl) | -4.7 [-9.2 to -0.2] | 0.04 |
| Sodium (mmol/L) | 1.0 [-0.4 to 2.3] | 0.15 |
| Potassium (mmol/L) | -0.3 [-0.5 to 0.0] | 0.025 |
| Magnesium (mmol/L) | 0.0 [-0.2 to 0.2] | 0.84 |
| Calcium (mmol/L) | 0.5 [0.3 to 0.7] | <0.001 |
| Phosphorous (mmol/L) | -0.1 [-0.4 to 0.2] | 0.56 |
| Uric acid (mg/dL) | 0.1 [-0.6 to 0.4] | 0.69 |
| NT-proBNP (pg/mL) | -1912 [-3117 to -708] | 0.002 |

\* Repeated measures ANOVA

**Figure 2:**
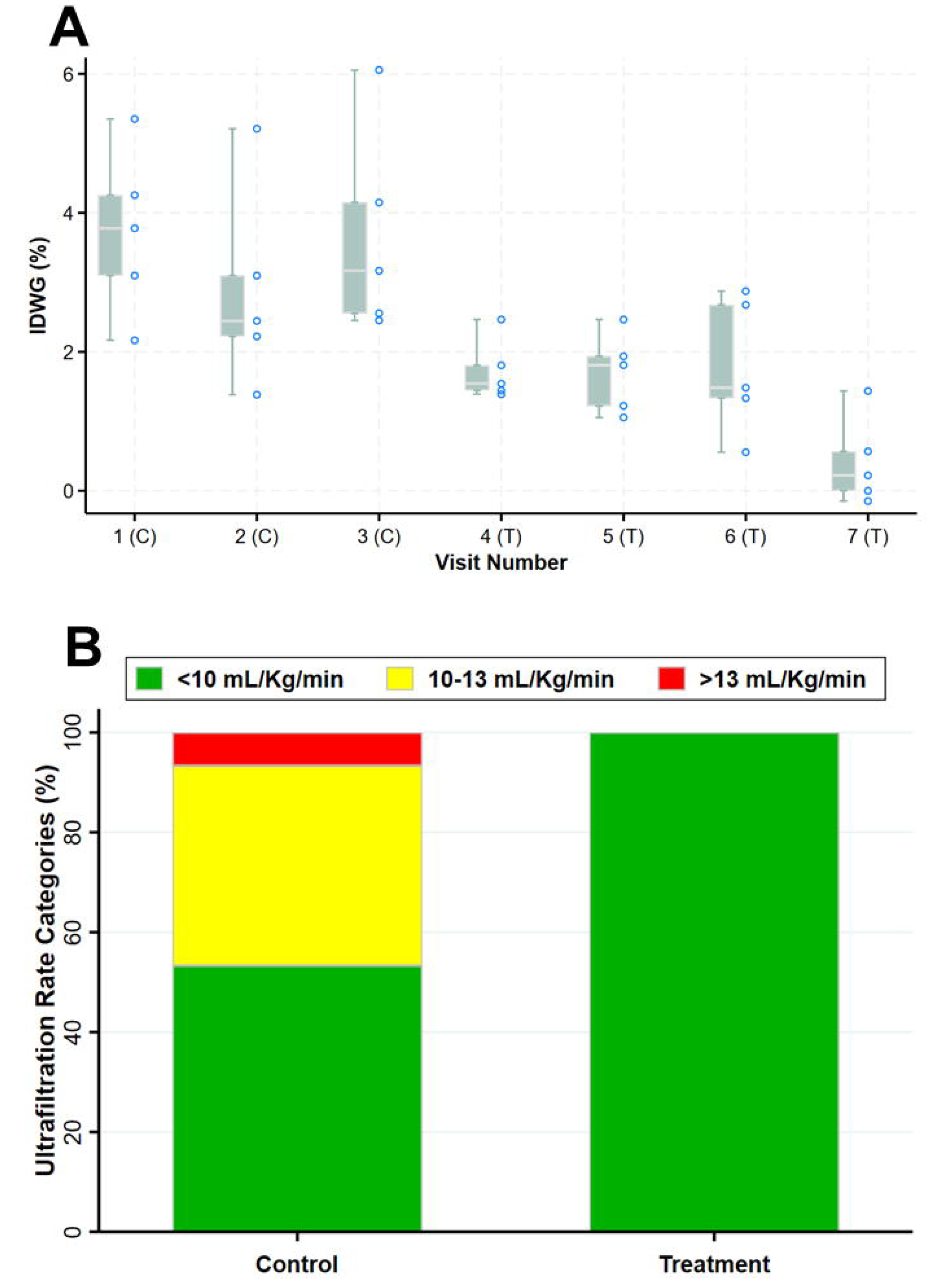
**A**. Box-and-whisker plot and scatter plot of IDWG% during the control (C) and treatment (T) phases. The line within the box denotes the median and the box spans the interquartile range (25–75^th^ percentiles). Whiskers extend from the 5^th^ to 95^th^ percentiles; **B**. Percent of patients in the 3 categories of ultrafiltration rate in the control and treatment phases.

**Figure 3:**
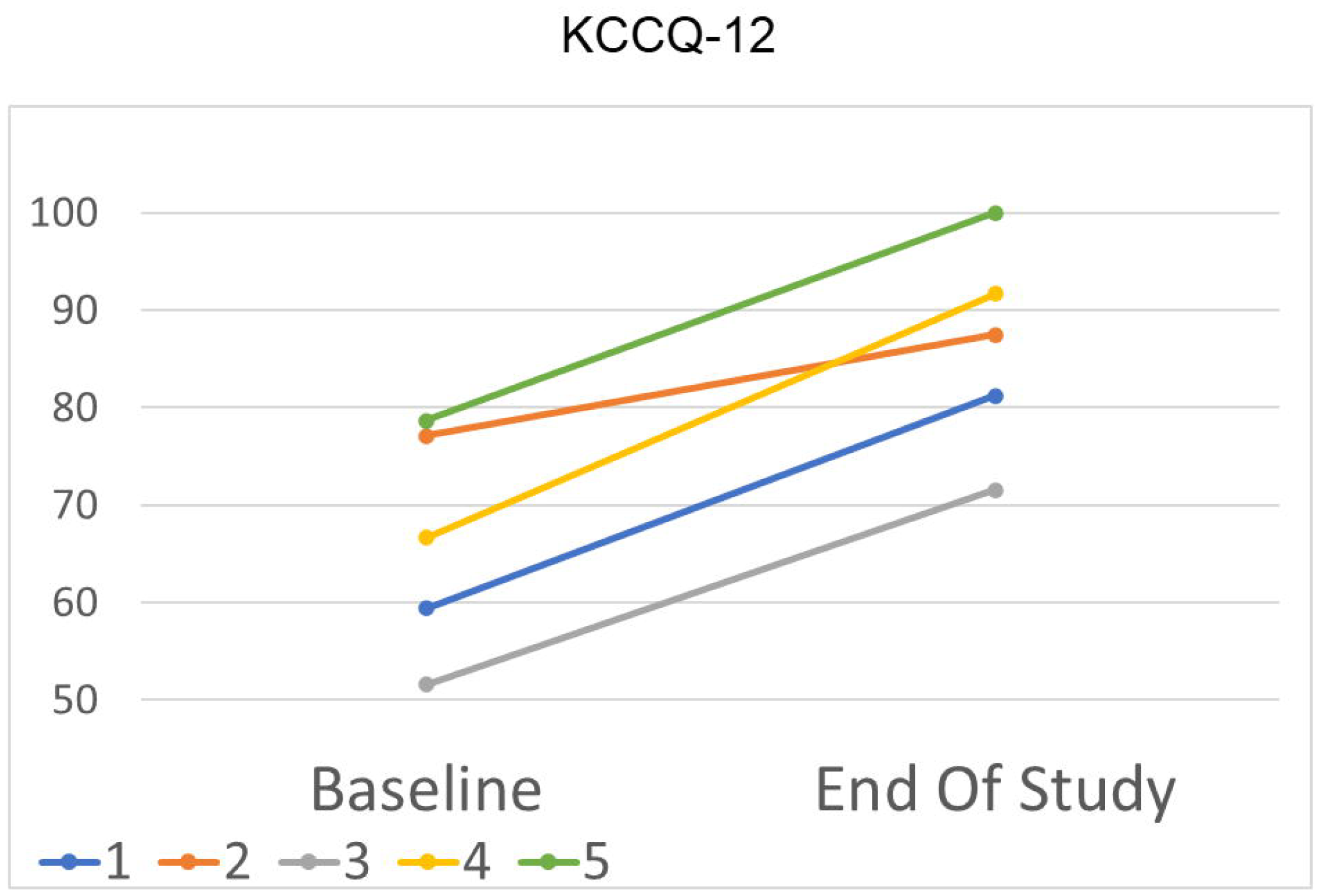
Change in quality of life as assessed by the Kansas City Cardiac Questionnaire-12 (KCCQ-12) during the study.

## Discussion

While achieving normovolemia is a central element of dialysis prescription, it is notoriously difficult to accomplish in clinical practice. The clinical hallmarks of high IDWG are diminished quality of life and repeated cycles of fluid overload leading to increased cardiovascular hospitalizations and increased mortality rates.^3^ Attempts to alleviate volume overload using dialysis time extension or additional dialysis sessions have been poorly accepted by patients. Therefore, high IDWG imposes more aggressive volume reduction during hemodialysis and high UF rate.

The study represents the first step in the clinical evaluation of a novel strategy to remove fluid by increasing sweat rate as a potential solution for IDWG. Fluid removal rates with the device were comparable to normal subjects and patients with chronic heart failure,^12^ and led to large reductions in IDWG thus enabling lower and safer UF rates. In addition, our preliminary data suggest a significant reduction in potassium levels. Given the observed safety and clinical performance of the device, this study supports further evaluation of this concept in HD patients.

With IDWG, the excess fluid initially redistributes into the extravascular space.^1, 15^ With increased sweat rate, the fluid is removed from the interstitial compartment because interstitial fluid is the precursor fluid for primary sweat. Removing fluids directly from the interstitial space therefore protects the intravascular compartment from underfilling prior to the subsequent hemodialysis.

In conclusion, the AquaPass device may provide important health benefits in hemodialysis patients by lowering IDWG and mitigating chronic volume overload present in many HD patients.^2^ Together with trice weekly HD, this therapy more closely approximates the capacity of the native kidney to regulate extracellular volume and solute composition, similar to patients with substantial RKF. This approach can also address long interdialytic gaps in resource-poor regions with limited access to thrice-weekly hemodialysis.

## Data Availability

All data produced in the present work are contained in the manuscrip

